# Towards a platform for robot-assisted minimally-supervised therapy of hand function: design and pilot usability evaluation

**DOI:** 10.1101/2021.01.12.21249685

**Authors:** Raffaele Ranzani, Lucas Eicher, Federica Viggiano, Bernadette Engelbrecht, Jeremia P.O. Held, Olivier Lambercy, Roger Gassert

## Abstract

**Background:** Robot-assisted therapy can increase therapy dose after stroke, which is often considered insufficient in clinical practice and after discharge, especially with respect to hand function. Thus far, there has been a focus on rather complex systems that require therapist supervision. To better exploit the potential of robot-assisted therapy, we propose a platform designed for minimal therapist supervision, and present the preliminary evaluation of its immediate usability, one of the main and frequently neglected challenges for real-world application. Such an approach could help increase therapy dose by allowing the training of multiple patients in parallel by a single therapist, as well as independent training in the clinic or at home.

**Methods:** We implemented design changes on a hand rehabilitation robot, considering aspects relevant to enabling minimally-supervised therapy, such as new physical/graphical user interfaces and two functional therapy exercises to train hand motor coordination, somatosensation and memory. Ten participants with chronic stroke assessed the usability of the platform and reported the perceived workload during a single therapy session with minimal supervision. The ability to independently use the platform was evaluated with a checklist.

**Results:** Participants were able to independently perform the therapy session after a short familiarization period, requiring assistance in only 13.46(7.69-19.23)% of the tasks. They assigned good-to-excellent scores on the SUS to the user-interface and the exercises (85.00(75.63-86.88) and 73.75(63.13-83.75) out of 100, respectively). Nine participants stated that they would use the platform frequently. Perceived workloads lay within desired workload bands. Object grasping with simultaneous control of forearm pronosupination and stiffness discrimination were identified as the most difficult tasks.

**Discussion:** Our findings demonstrate that a robot-assisted therapy device can be rendered safely and intuitively usable upon first exposure with minimal supervision through compliance with usability and perceived workload requirements. The preliminary usability evaluation identified usability challenges that should be solved to allow real-world minimally-supervised use. Such a platformcould complement conventional therapy, allowing to provide increased dose with the available resources, and establish a continuum of care that progressively increases therapy lead of the patient from the clinic to the home.

## 1 Introduction

Despite progress in the field of neurorehabilitation over the last decades, around one third of stroke survivors suffer from chronic arm and hand impairments (Raghavan, 2007; Morris et al., 2013), which limit their ability to perform basic activities of daily living (Jönsson et al., 2014; Franck et al., 2017; Katan and Luft, 2018). Growing evidence suggests that intensive rehabilitation programs that maximize and maintain therapy dose (i.e., number of exercise task repetitions and total therapy time) may promote further recovery and maintenance of upper limb function (Lohse et al., 2014; Veerbeek et al., 2014; Lang et al., 2015; Schneider et al., 2016; Vloothuis et al., 2016; Ward et al., 2019). A promising approach to increase therapy dose and intensity, may be offered by robot-assisted therapy (McCabe et al., 2015; Veerbeek et al., 2017; Daly et al., 2019), which has often been proposed as a complement to conventional physical therapies (Chang and Kim, 2013; Lambercy et al., 2018; Grosmaire et al., 2019). However, robot-assisted therapy has so far typically been applied in the context of short (frequently outpatient) therapy sessions in clinical settings (Lum et al., 2012; Page et al., 2013; Klamroth-Marganska et al., 2014), and where the presence of a supervising therapist is required to prepare and manage the complex equipment, set up the patient on/in the device and configure the appropriate therapy plan. This generates organizational and economical constraints that restrict the use of the technology (Wagner et al., 2011; Schneider et al., 2016; Rodgers et al., 2019; Ward et al., 2019) and, as a result, despite claiming high intensity (Lo et al., 2010; Rodgers et al., 2019), the therapy dose achieved using robots remains limited compared to guidelines (Bernhardt et al., 2019) and preclinical evidence (Nudo and Milliken, 1996).

Minimally-supervised therapy, defined here as any form of therapy performed by a patient independently with minimal external intervention or supervision, is a promising approach to better harvest the potential of rehabilitation technologies such as robot-assisted therapy (Ranzani et al., 2020). This could allow for the simultaneous training of multiple subjects in the clinics (Büsching et al., 2018), or for subjects to receive robot-assisted training in their home (Chi et al., 2020). Several upper limb technology-supported therapies have been proposed for home use (Wittmann et al., 2016; Ates et al., 2017; Nijenhuis et al., 2017; Chen et al., 2019; Cramer et al., 2019; Laver et al., 2020), allowing subjects to benefit from additional rehabilitative services to increase dose (Laver et al., 2020; Skirven et al., 2020). However, only few minimally-supervised robotic devices capable of actively supporting/resisting subjects during interactive therapy exercises have been proposed (Lemmens et al., 2014; Sivan et al., 2014; Wolf et al., 2015; Hyakutake et al., 2019; McCabe et al., 2019) and, as typically happening in conventional care (Qiuyang et al., 2019), most of them did not focus on the hand. Moreover, these devices only partially fulfilled the complex set of constraints imposed by a minimally-supervised use.

To be effective, motivating and feasible, minimally-supervised robot-assisted therapy platforms (i.e. a set of hardware and software technologies used to perform therapy exercises) should meet a wide range of usability, human factors and hardware requirements, which are difficult to respect simultaneously. Besides the necessity to provide motivating and physiologically relevant task-oriented exercises to maximize subject engagement (Veerbeek et al., 2014; Laut et al., 2015; French et al., 2016; Johnson et al., 2020) and to monitor subjects’ ability level to continuously adapt the therapy (Metzger et al., 2014a; Hocine et al., 2015; Wittmann et al., 2015; Aminov et al., 2018), ensuring ease of use is critical (Zajc and Russold, 2019). When considering using a robot-assisted platform in a minimally-supervised way (clinical and home settings), specific hardware and software changes should be considered to allow a positive user experience and compliance with the therapy program/targets, as well as to assure safe interaction. In this sense, integrating usability evaluation during the development of rehabilitation technologies was shown to contribute to device design improvements, user satisfaction and device usability (Shah and Robinson, 2007; Power et al., 2018; Meyer et al., 2019). Unfortunately, the usability of robotic devices for upper-limb rehabilitation is only rarely evaluated and documented in the target user population before clinical tests (Pei et al., 2017; Catalan et al., 2018; Guneysu Ozgur et al., 2018; Nam et al., 2019; Tsai et al., 2019).

In this paper, we present the design of a platform for minimally-supervised robot-assisted therapy of hand function after stroke and the evaluation of its short-term usability in a single experimental session with 10 potential users in the chronic stage after stroke. The proposed platform builds on an existing high-fidelity 2-degrees-of-freedom end-effector haptic device (ReHapticKnob (Metzger et al., 2014b)). This device was successfully applied in a clinical trial on subjects with subacute stroke, showing equivalent therapy outcomes in a supervised clinical setting compared to dose-matched conventional therapy without any related adverse event (Ranzani et al., 2020). This was a prerequisite for the exploration of strategies to better take advantage of the robot’s unique features, such as potentially allowing the provision of minimally-supervised therapy. In order to make the device usable in a minimally-supervise setting, we developed an intuitive user interface (i.e., physical/hardware and graphical/software) that can be independently used by subjects in the chronic stage after stroke to perform therapy exercises with minimal supervision, either in clinical settings or at home. Additionally, two new task-oriented therapy exercises were developed to be used in a minimally-supervised scenario and complement existing exercises by simultaneously training hand grasping and forearm pronosupination, which are functionally relevant movements. The goal of this paper is to present the hard and software modifications to the platform (i.e., robotic device with new physical and virtual user interface and therapy exercises) as well as the results of a preliminary usability evaluation with participants in the chronic stage after stroke using the device independently in a single session after a short explanation/familiarization period. This work is an important step to demonstrate that subjects with chronic stroke can independently and safely use a powered robotic device for upper-limb therapy upon first exposure, highlight key design aspects that should be taken into account for maximizing usability in real-world minimally-supervised scenarios, and thereby provide a methodological basis that could be generalized to other platforms and applications.

## 2 Materials and Methods

### 2.1 ReHapticKnob and User Interface

The therapy platform (hardware and software) proposed in this work consists of an existing robotic device, the ReHapticKnob (RHK, (Metzger et al. 2011)) with a new user interface and two novel therapy exercises. The user interface is assumed to include physical components (i.e., hardware interfaces that get in contact with the user, such as the finger pads and buttons of the rehabilitation device, the ReHapticKnob) and a graphical component (i.e. software), i.e. the graphical user interface (GUI).

The RHK is a 2-degrees-of-freedom haptic device for assessment and therapy of hand function after stroke. It incorporates a set of automated assessments to determine the baseline difficulty of the therapy exercises (Metzger et al., 2014b), and allows to train hand opening-closing (i.e., grasping) and pronosupination of the forearm by rendering functionally-relevant rehabilitative tasks (e.g., interaction with virtual objects) with high haptic fidelity (Metzger et al., 2014a). The user sits in front of the robot positioning his/her hand inside two instrumented finger pads with VELCRO straps, as shown in Figure 1. The simple end-effector design of the robot (compared to the typically more complex donning and doffing of exoskeletons where joint alignment is critical) makes it an ideal candidate for independent use.

**Figure 1.**
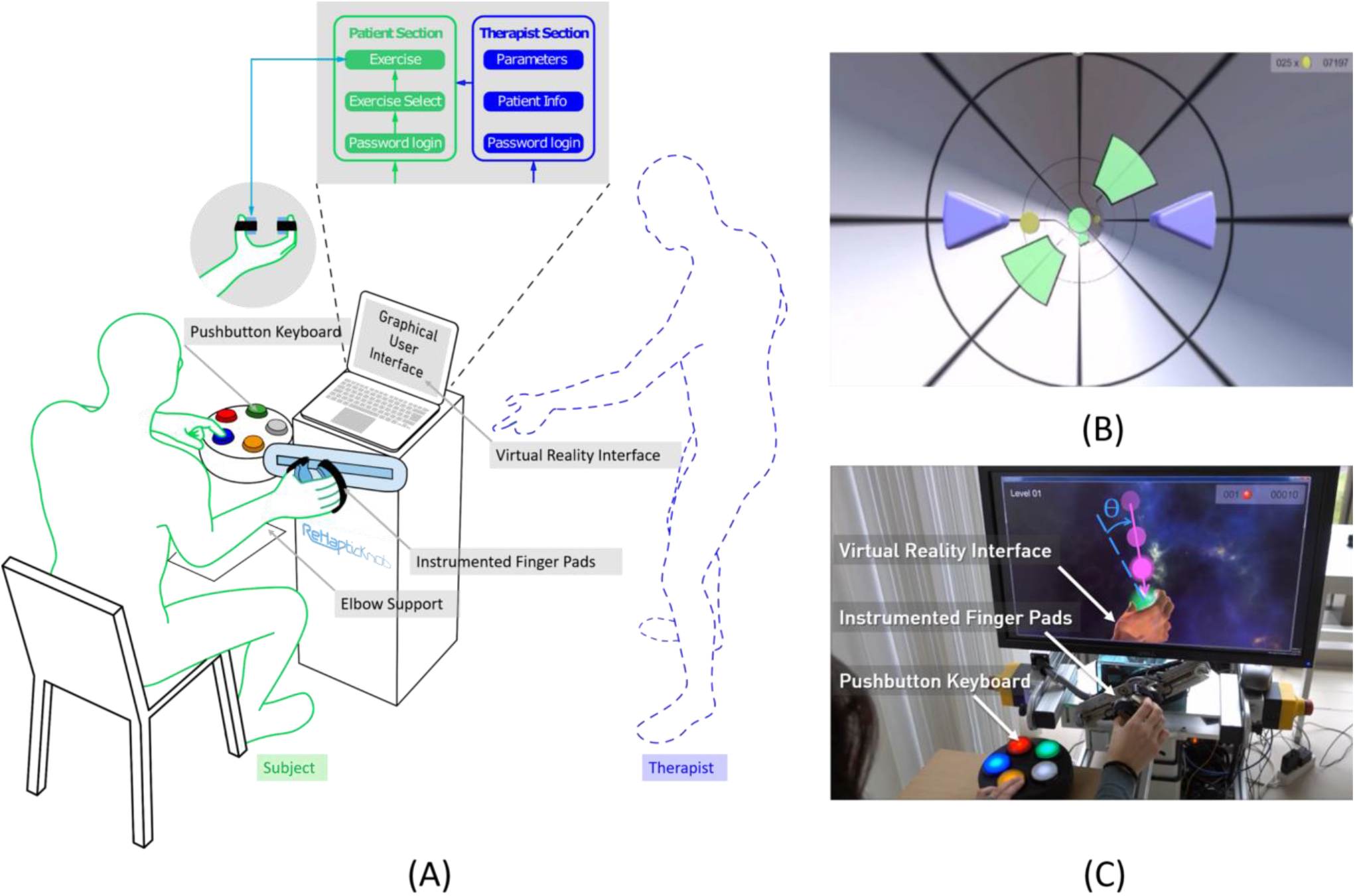
The ReHapticKnob therapy platform. (A) The platform consists of a haptic rehabilitation device - the ReHapticKnob - with physical (i.e., instrumented finger pads, colored pushbutton keyboard) and graphical user interfaces and a set of therapy exercises that can be used with minimal supervision. The graphical user interface includes a section for the therapist to initially customize the therapy plan and a patient section through which the user can autonomously perform predefined therapy exercises. (B) Virtual reality interface of the tunnel exercise. The subject has to drive a set of purple avatars by opening-closing and pronosupinating the finger pads. The goal is to avoid the green obstacles and collect as many coins as possible. (C) A subject performing the sphere exercise on the ReHapticKnob. During the testing phase shown, the subject has to catch a falling sphere halo by rotating the finger pads (pronosupination). The object is caught if the hand orientation (dotted line) is aligned with the falling direction (continuous line), within a certain angular range ϴ. Once the object is caught, the subject selects the sphere stiffness he/she perceives while squeezing the object by pressing the corresponding color on the pushbutton keyboard.

Considering the feedback collected from subjects and therapists within a previous clinical study under therapist supervision (Ranzani et al., 2020), we embedded the RHK into a novel therapy platform that is more user-friendly and suitable for minimally-supervised use. For this purpose, a novel GUI now directly controls the execution of the therapy program and includes two sections, one for the user and one for the therapist to customize the therapy, for example, before the first therapy session (Figure 1A). To configure the therapy for a subject, the therapist can log into a password protected “Therapist Section”, create/update a subject profile (i.e., selected demographic data, impaired side, identification code and password consisting of a sequence of 4 colors to access the therapy plan) and select the relevant exercise parameters (i.e., derived from preliminary automated assessments) that are needed to adapt the therapy exercises to the subject ability level (Metzger et al., 2014b). To perform a therapy session, the subject can autonomously navigate into the “Patient Section” using an intuitive colored pushbutton keyboard (Figure 1A). The subject can log in into his/her therapy plan by selecting his/her identification code and typing the defined colored password on the pushbutton interface. In the personal therapy plan, a graphical list of all the available therapy exercises appears. The user can then manually navigate through the exercise list and select the preferred exercise.

To maximize the usability and, consequently, the likeliness of therapists and subjects using the device, attention was devoted to the optimization of esthetics and simplicity in all virtual displays and hardware components. The design was guided by a set of usability heuristics (Nielsen, 1995), which included visibility of feedbacks (e.g., show performance feedback and unique identifiers on each user interface window), matching between virtual and haptic displays of the platform and corresponding real world tasks, user control and freedom (e.g., exit or stop buttons always available), consistency and standardization of displays’ appearance, visibility and intelligibility of instructions for use of the system, fast system response, pleasant and minimalistic design, as well as simple error detection/warnings. Particular caution was directed to the placement (e.g., easily visible/retrievable), size (e.g., large to be easily selectable), logical ordering, appearance and color coding (e.g., red for quitting/exiting) of the buttons both in the colored pushbutton keyboard and in the virtual displays (Norman, 2013; de Leon et al., 2020), trying to reduce them to a maximum of five, which was needed to execute all exercises. Finally, to guarantee platform modularity, a state machine performs the low-level control of the robot (i.e., position, velocity and force control implemented in LabVIEW 2016) while the graphical user interface (Unity 5.6) guides the high-level control of the therapy session and easily allows to insert/remove different exercise types.

### 2.2 Exercises for Robot-Assisted Minimally-Supervised Therapy

The RHK includes a set of seven assessment-driven therapy exercises (Metzger et al., 2014b), which were developed following the neurocognitive therapy approach formulated by Perfetti (i.e., combining motor training with somatosensory and cognitive tasks) (Perfetti and Grimaldi, 1979). So far, these exercises only focused on the training of isolated movements (i.e., grasping or pronosupination) and were administered under therapist supervision (see (Metzger et al., 2014b; Ranzani et al., 2020) for more details on existing exercises).

To complement this available set of exercises, we implemented two new exercises optimized for use in a minimally-supervised scenario and focusing on tasks that should facilitate a transition to activities of daily living. For this purpose, the exercise tasks train synchronous movements and combine complex elaborations of sensory, cognitive and motor cues.

The tunnel exercise is a functional exercise focusing on synchronous coordination of grasping and pronosupination and on sensory perception of haptic cues. The user has to move two symmetric avatars (virtually representing the finger pads of the robot as two purple triangles, see Figure 1B) progressing in a virtual tunnel, while avoiding obstacles and trying to collect as many rewards/points (e.g., coins) as possible. The exercise includes sensory cues, namely hand vibrations indicating the correct position to avoid an obstacle, stiff virtual walls that constrain the movement of the avatars inside the virtual tunnel, and changes in viscosity (i.e., velocity-dependent resistance) within the tunnel environment on both degrees of freedom to challenge the stabilization of the hand movement during navigation. Increasing difficulty levels linearly increase the avatars speed within the tunnel (while consecutive obstacles remain at a constant distance with respect to one other), the maximum pronation and supination locations of the apertures between obstacles to promote an increase in the pronosupination range of motion (ROM) of the subject (based on an initial robotic assessment) and the changes in environment viscosity, while the space to pass through the obstacles and the haptic vibration intensity are linearly decreased. One exercise block consists of a one-minute long progression within the virtual tunnel, where up to 30 obstacles have to be avoided. One exercise session consists of a series of ten one-minute blocks.

The sphere exercise is a functional exercise focusing on hand coordination during grasping and pronosupination, with a strong focus on somatosensation and memory to identify the objects that are caught. One exercise block consists of a training phase and a testing phase. In the training phase, the user moves a virtual hand and squeezes a set of virtual spheres (i.e., three to five) to memorize the color attributed to each stiffness rendered by the robot (for more details refer to (Ranzani et al., 2019)). The user can manually switch the sphere to try/squeeze by pressing a predefined button on the colored pushbutton keyboard. In the testing phase, semi-transparent spheres (halos) fall radially from a random initial position, one at a time, towards the hand. By actively rotating the robot and adjusting the hand opening, the user has to catch the falling halo. A halo is only caught if the hand aperture matches the sphere diameter within an error band of ±10mm, and the hand pronosupination angle is aligned with the falling direction within an error band Ɵ (see Figure 1C) between ±40° and ±15° depending on the difficulty level. When a halo is caught, the participant has to squeeze it, identify its stiffness, and indicate it using the colored pushbutton keyboard. Each testing phase lasts 3 min. At increasing difficulty levels, the number of spheres and the speed of the falling halos increase, the tolerance in hand positioning to grasp the falling halos are reduced in the pronosupination degree of freedom, and the relative change in object stiffness decreases as a function of the subject’s stiffness discrimination ability level (based on initial robotic psychophysical assessments). One exercise session consists of three blocks (i.e., three training phases, each followed by a testing phase) and lasts between 10 and 15 minutes.

In both exercises, at the end of a block, the achieved performance (i.e., percentage of obstacles avoided over total obstacles for the tunnel exercise, and the percentage of spheres correctly caught and identified over total number of halos for the sphere exercise) is summarized to the subject through a score display before a new block begins. The assessment-driven tailoring of the exercise difficulty from the first therapy session should allow the subject to maintain a performance around 70%, which maximizes engagement and avoids the frustration that would arise when performance is too low or too high (Adamovich et al., 2009; Cameirão et al., 2010; Choi et al., 2011; Lambercy et al., 2011; Metzger et al., 2014b; Wittmann et al., 2015).

### 2.3 Participants

A pilot study to evaluate the usability of the proposed minimally-supervised therapy platform was conducted on ten subjects with chronic stroke (>6 months), representative of potential future users of the platform. Subjects were enrolled if they were above 18 years old, able to lift the arm against gravity, had residual ability to flex and extend the fingers, and were capable of giving informed consent and understanding two-stage commands. Subjects with clinically significant non-related pathologies (i.e., severe aphasia, severe cognitive deficits, severe pain), contraindications on ethical grounds, known or suspected non-compliance (e.g., drug or alcohol abuse) were excluded from the study.

### 2.4 Pilot Study Design

The pilot study was conducted at ETH Zurich, Switzerland, over a period of two weeks. Participants took part in a single test session, as illustrated in Figure 2. The session consisted of a supervised and a minimally-supervised part. In the supervised part, a supervising professional therapist assessed the subject’s baseline ability level through a set of standard clinical and robotic assessments, which were used to customize the difficulty levels of the therapy exercises. The therapist then instructed the subject on how to perform the exercises, and actively guided the subject in the execution of one block of each exercise. In this part of the experiment, subjects were encouraged to ask any questions they had related to the use of the device. In the subsequent minimally-supervised part, the subject had to independently use the therapy platform to perform the tunnel exercise (10 blocks) and the sphere exercise (3 blocks). During that time, the therapist sat at the back of the room and silently observed the subject’s actions, recording any error or action that the subject could not perform in a checklist, and intervening only in case of risk or explicit request from the subject. The subject had to independently place his/her hand inside the finger pads, log into the therapy plan (i.e., find his/her identification code through other subject identification codes and insert the personal colored password to log in), find and start the appropriate therapy exercises from a list of all available RHK exercises, test both exercises and log out from the therapy plan. At the end of the experimental phase, subjects answered a set of usability questionnaires. The study was approved by the Cantonal Ethics Committee in Zurich, Switzerland (Req-2017-00642).

**Figure 2.**
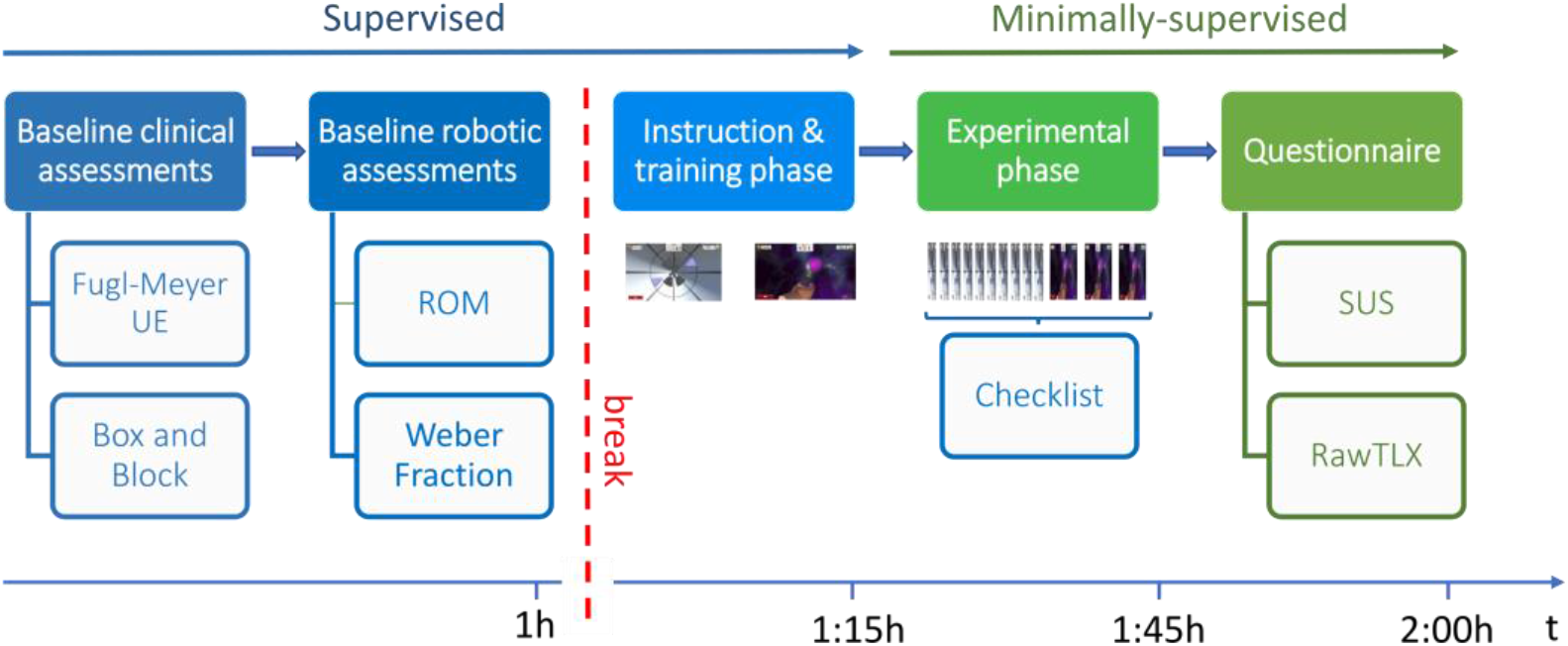
Study protocol. Abbreviations: UE – Upper Extremity. ROM – Range of Motion. SUS – System Usability Scale. RawTLX – Raw Task Load Index.

### 2.5 Baseline Assessments

Subjects’ upper limb impairment was measured at baseline with the Fugl-Meyer Assessment of the Upper Extremity (FMA-UE) (Fugl-Meyer et al., 1975) and its wrist and hand subscore (FMA-WH). Gross manual dexterity was assessed using the Box and Block test (BBT) (Mathiowetz et al., 1985). In addition to clinical assessments, robotic assessments (i.e., ROM and stiffness discrimination ability expressed as Weber fraction) were performed and used to adapt the initial difficulty level of the therapy exercises to the subject’s ability from the first block of the exercise. “ROM” assesses the subject’s ability to actively open and close the hand and pronosupinate the forearm. In the tunnel game this scales the positioning and size of the virtual walls that determine the size of the tunnel, while in the sphere exercise, this scales the workspace of the falling halos. “Weber fraction” describes the smallest distinguishable difference between two object stiffnesses and scales the initial stiffness difference between spheres in the sphere exercise. For more details on the robotic assessments, please refer to (Metzger et al., 2014b).

### 2.6 Outcome Measures and Statistics

To evaluate the ability of chronic stroke subjects to independently use the therapy platform and identify remaining usability challenges, the main outcome measures of this study were a performance checklist to record the tasks/actions that the subject could, or could not perform without supervision or in which therapist help was required, followed by two standardized usability questionnaires. To evaluate each component of the therapy platform separately, the performance checklist and the two questionnaires were repeated for the user interfaces (i.e., GUI and hardware interfaces such as the finger pads and pushbutton keyboard) and for the two exercises. The performance checklist includes 26 items, grouped as follows. Seven items about the use of the user interface verified if the subject could place/keep the hand inside the device (U1), use the pushbutton to navigate to his/her identification code (U2) and select it (U3), insert his/her color-code password to log into the therapy plan (U4), navigate to the therapy exercise (U5) and run it (U6), and quit the exercise at any time (U7). Six items related to the tunnel exercise verified if the subject could close the finger pads to a minimum position for calibration (TU1), open the finger pads to a comfortable position for playing the exercise (TU2), start the exercise (TU3), play one exercise block (TU4), start the next block once ready (TU5), and carry out ten exercise blocks (TU6). Thirteen items related to the sphere exercise verified if the subject could put the handlebar in a horizontal position (SP1) and close the finger pads to a minimum position for calibration (SP2), start the exercise (SP3), maintain the hand relaxed in a neutral position (SP4) after squeezing each available sphere in the training phase (SP5), repeat the training phase (SP6) or start the testing phase (SP7) when needed, open the finger pads and align the handlebar and catch the halo (SP8), squeeze the halo (SP9) and identify its stiffness (SP10), carry out one exercise block (SP11), start the next block once ready (SP12), and perform three exercise blocks (SP13).

The standardized usability questionnaires were:

System Usability Scale (SUS) (Brooke, 1996), a ten-item questionnaire which assesses the overall usability (i.e., effectiveness, efficiency, satisfaction) of the system under investigation (i.e., user interface and each of the two exercises separately). Two items of the SUS refer specifically to the “learnability” of a system (i.e., “I think that I would need the support of a technical person to be able to use this system”, “I needed to learn a lot of things before I could get going with this system”) and were considered of high importance for the evaluation of the minimally-supervised usage scenario (Lewis and Sauro, 2009). Ideally, the total SUS score calculated from its ten items should be greater than 50 out of 100, indicating an overall usability between “OK” and “best imaginable” (Bangor et al., 2009). To evaluate if there was any correlation between SUS score and baseline characteristics of the subjects, we calculated the Pearson correlation between the SUS score and age, FMA-UE, FMA-WH, and BBT. The statistical significance level of α = 0.05 was corrected using Bonferroni correction, leading to a value of 0.0042.

Raw Task Load Index (RawTLX) (Hart, 2006), a six-item questionnaire which assesses the workload while using the system under investigation. The RawTLX is the widely used, shortened form of the original NASA TLX, with the difference of the six workload domains being evaluated individually without the calculation of a total workload score through domain-weighting. The workload domains assessed are: (i) mental demand (i.e., amount of mental or perceptual activity required), (ii) physical demand (e.g., amount of physical workload required), (iii) temporal demand (i.e., amount of time pressure perceived during the use), (iv) overall performance (i.e., perceived level of unsuccessful performance), (v) effort (i.e., total amount of effort perceived to execute the task), and (vi) frustration level (i.e., amount of stress/irritation/discouragement perceived). The target workload levels differ depending on the application, thus they were defined by the investigator and therapist. For the user interface, a targeted minimal workload (i.e. ≤25%) was set as goal in all domains except for temporal demand, in which an intermediate workload level (i.e., between 25% and 75% included) was tolerated. The exercises should be challenging but not too difficult (Adamovich et al., 2009; Choi et al., 2011), allowing the subjects to maintain actual and perceived performance around 70%. For this reason, a target workload between 50 and 75% (included) was desired for mental, physical, temporal and effort domains, and a corresponding workload between 25% and 50% was desired in the performance domain, in which the workload axis is inversely proportional to the perceived performance. Finally, frustration should be avoided, so a workload ≤25% is required.

The SUS and RawTLX questionnaires were translated to German by a native speaker and rated on a 5-intervals Likert scale, which was associated with corresponding scores of 0, 25, 50, 75, and 100%.

To monitor the safety of the platform during the minimally-supervised part of the experiment, adverse events and situations that could put at risk the safety of the user (e.g., triggering of safety routines of the robot for excessive forces/movements or hardware/software errors) were recorded.

Given the relatively small sample size and the non-normal distribution of the majority of the data presented in the paper, results are reported as median with first and third quartile (i.e., median(Q1-Q3)).

## 3 Results

### 3.1 Experiment Characteristics

Ten subjects (4 female, 6 male) in the chronic stage after an ischemic stroke (39.50(27.00-60.50) months post event) were eligible and agreed to participate in the study. The participant age was 60.50(56.25-67.50) and there were 4 right and 6 left hemisphere lesions, while all subjects were right-handed. Most subjects showed mild to moderate (Woytowicz et al., 2017) initial upper-limb impairment with a FMA-UE of 41.50(39.25-50.00) out of 66 points, and a FMA-WH of 17.00(14.00-19.50) out of 24 points. In the BBT, subjects transported 39.50(30.00-48.75) blocks in one minute using their impaired limb. Before enrollment, all participants were informed about the study and gave written consent.

The experiment lasted 111.50(104.00-135.00) minutes, which included 79.12(67.00-86.00) minutes of robot use and 34.75(24.00-48.00) minutes for baseline clinical assessments, break time and questionnaires. Within the robot use, the subjects spent 16.00(14.00-20.00) minutes on baseline robotic assessments and 59.93(53.00-67.00) minutes to learn how to use the user interface and exercises (i.e., instruction and training phase, 27.56(22.00-38.00) minutes) and test them with minimal supervision (i.e., experimental phase, 30.63(28.00-32.00) minutes). During the experimental phase, the therapist’s physical intervention (e.g., to assist hand movements or position the hand) or suggestions and further explanations (e.g., to repeat the login password or refresh the exercise rules) were required 3.50(2.00-5.00) times per subject out of the 26 checklist items (see Figure 3), with highest number of interventions required by the oldest subject (subject 3, 87 years old, 7 interventions).

**Figure 3.**
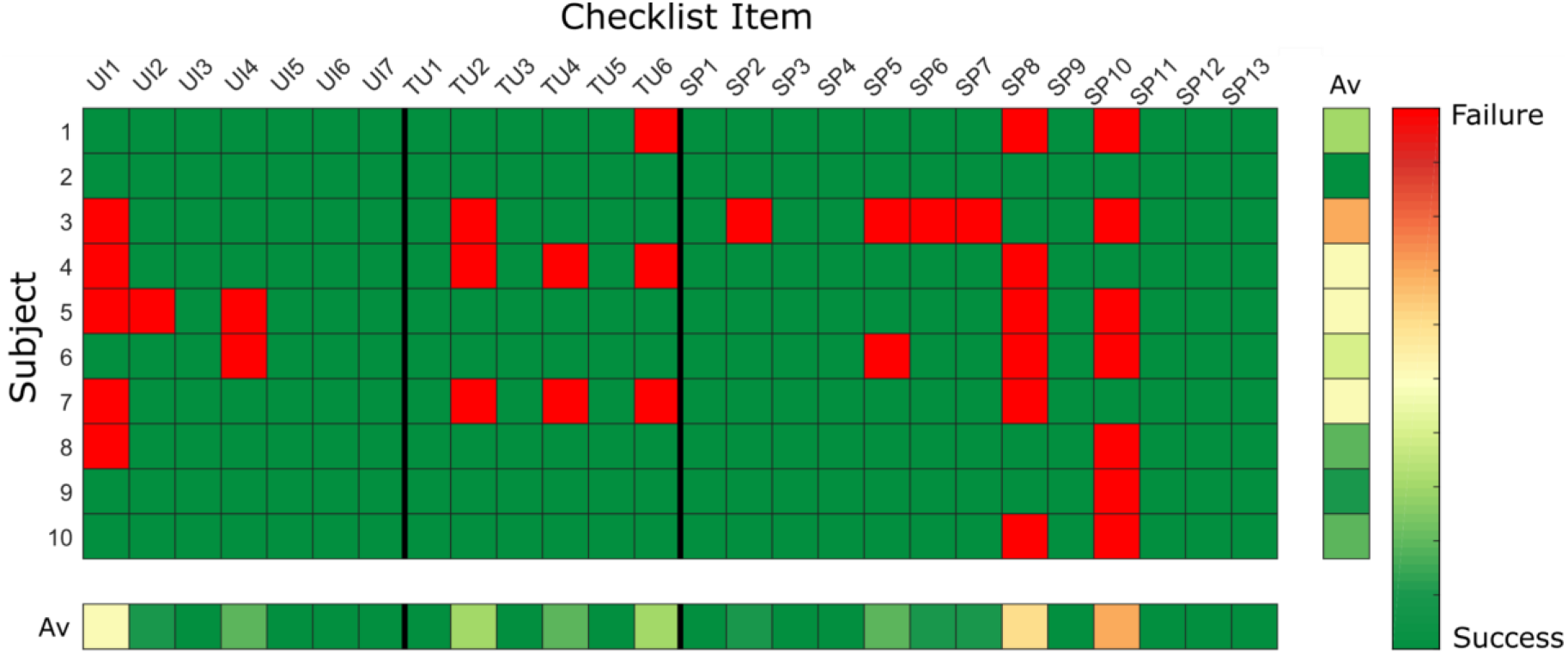
Checklist results represented as heatmap. The results averaged over subjects and items are presented on the bottom and on the right of the heat map, respectively. (Green: no problem/issue in item completion without external intervention; Red: Failure and/or external intervention required to solve the item; Av: average).

Over the duration of the study, no serious adverse event related to the robot-assisted intervention or event that would put at risk the safety of the user were observed, but the software had to be restarted two times due to the triggering of safety routines (e.g., too high forces, positions, velocities generated by the user). Two subjects reported a mild temporary increase in hand muscle tone (e.g., finger flexors and/or extensors) during the therapy exercises.

### 3.2 User Interface

The user interface was ranked with a SUS score between good and excellent (85.00(75.63-86.88) out of 100) as shown in Figure 4A, and a learnability score of 15.00(13.13-16.88) out of 20. Nine subjects gave excellent rating and reported that they would use the RHK frequently. Most of the subjects reported that the user interface is intuitive and that the colored button interfaces are easy to use. The oldest subject (age 87) gave a score in the region of “worst imaginable” for the user interface (as well as for the sphere and tunnel exercises). The SUS results showed an inverse relationship with the age of the subjects, but no significant correlation following Bonferroni correction (correlation −0.737, p-value 0.015), and no linear relationship with their ability level as measured with the FMA-UE (0.170, 0.639), FMA-WH (-0.044, 0.904), and BBT (0.207, 0.566) scales.

**Figure 4.**
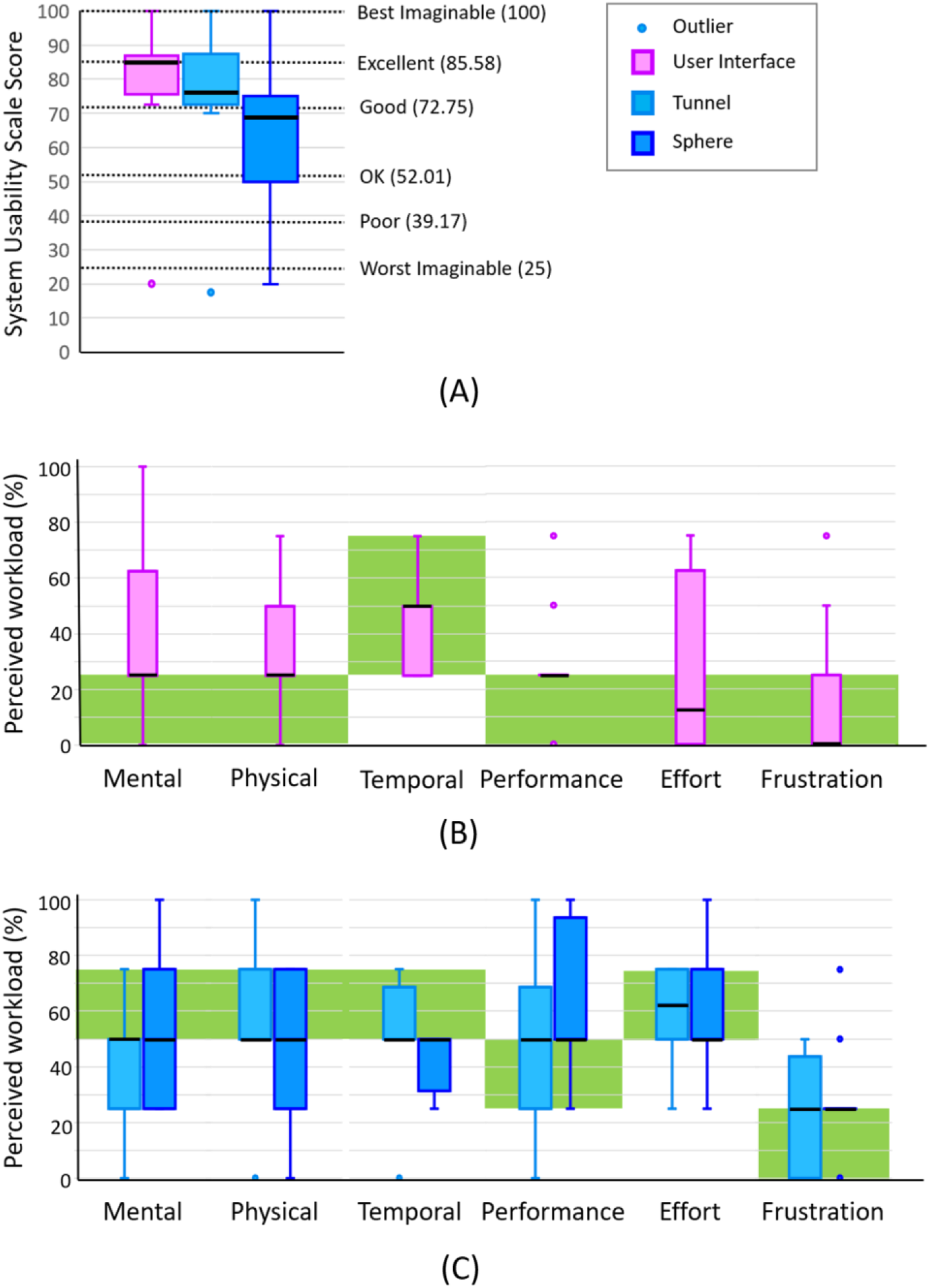
(A) System Usability Scale results for user interface (i.e., GUI, finger pads and pushbutton keyboard), tunnel exercise and sphere exercise. (B) Raw TLX perceived workload levels for user interface and (C) for tunnel and sphere exercise. black line: median; green area: target workload level.

The Raw TLX results are shown in Figure 4B. The median perceived workload levels lie within the target workload band for mental demand (25.00(25.00-62.50)%), physical demand (25.00(25.00-50.00)%), temporal demand (50.00(25.00-50.00)%), performance (25.00(25.00-25.00)%), effort (12.50(0.00-62.50)%) and frustration (0.00(0.00-25.00)%). However, the third quartile is outside of the target band (higher) for at least one datapoint (25%) in mental demand, physical demand and effort.

The subjects required external supervision or assistance for 14.29(0.00-14.29)% of the checklist items related to the user interface (Figure 3). Five subjects needed help to insert or reinsert the hand into the finger pads, as the thumb can easily slip out while moving the finger pads, particularly during the execution of active tasks within the exercises. Two subjects could not remember the colored password.

### 3.3 Tunnel Exercise

The usability of the tunnel exercise was ranked between good and excellent (76.25(72.50-87.50) out of 100) on the SUS (Figure 4A), while the learnability of the exercise was ranked 15.00(10.00-19.38) out of 20. Three subjects reported that the exercise is entertaining and motivating. As for the user interface, the SUS scores showed an inverse relationship with the age of the subjects, although without significant correlation after Bonferroni correction (correlation −0.681, p-value 0.030), and no linear relationship with FMA-UE (0.342, 0.333), FMA-WH (0.019, 0.958), and BBT (0.335, 0.344) scores.

The Raw TLX results are shown in Figure 4C. The median perceived workload levels lie within the target workload band for mental demand (50.00(25.00-50.00)%), physical demand (50.00(50.00-75.00)%), temporal demand (50.00(50.00-68.75)), performance (50.00(25.00-68.75)%), effort (62.50(50.00-75.00)%) and frustration (25.00(0.00-43.75)%). However, the first quartile is lower than the target workload band for at least one datapoint (25%) in mental demand.

The subjects required external supervision or assistance in only 0.00(0.00-16.67)% of the checklist items related to the tunnel exercise (Figure 3). Six out of ten subjects could perform the entire exercise independently without any therapist intervention. One subject could not independently perform the calibration at the beginning of the exercise as she did not understand the instructions provided by the robot (i.e., the robot was asking to open the hand to a comfortable position and the subject tried to open the hand as much as possible). Two subjects could not independently perform either the calibration or the hand opening/closing tasks of the exercise, as they could not actively open the hand beyond of the minimum position of the robot (i.e., approximately 4cm between thumb and index finger tip) due to their motor impairment level (i.e., FMA-UE below 38 out of 66 points, FMA-WH below 15 out of 24 points). One subject only completed 8 out of 10 blocks, as the robot went into a safety stop (i.e., too high forces, position or velocity). Two subjects required further explanations of the scope and rules of the exercise (e.g., tried to hit the obstacles instead of avoiding them). As additional comments, one subject reported that the tunnel speed was too fast for her, and another subject reported that the depth perception of the virtual reality should be improved. The median performance (i.e., number of obstacles avoided versus total number of obstacles) of the subjects within the ten blocks was 71.73(59.06-79.92)%, which is very close to the desired 70% performance.

### 3.4 Sphere Exercise

The usability of the sphere exercise was ranked between OK and good (68.75(50.00-75.00) out of 100) at the SUS (Figure 4A), while its learnability was ranked 10.00(10.00-14.38) out of 20. Two subjects reported that the game was too challenging and more boring compared to the tunnel exercise and would recommend this game for a mildly impaired population. As for the user interface and tunnel exercise, the SUS scores showed an inverse relationship with the age of the subjects without a significant correlation after Bonferroni correction (correlation −0.739, p-value 0.015), and no linear relationship with FMA-UE (0.092, 0.800), FMA-WH (0.014, 0.970), and BBT (0.081, 0.825).

The Raw TLX results are shown in dark blue in Figure 4C. The median perceived workload levels lie within the target workload band for mental demand (50.00(25.00-75.00)%), physical demand (50.00(25.00-75.00)%), temporal demand (50.00(31.25-50.00)), performance (50.00(50.00-93.75)%),effort (50.00(50.00-75.00)%) and frustration (25.00(25.00-25.00)%). However, the first quartile is lower than the target workload band for at least one datapoint (25%) in mental and physical demand, and the third quartile is higher than the target workload band in performance.

The subjects required external supervision or assistance in 11.54(7.69-15.39)% of the checklist items related to the sphere exercise (Figure 3). Only one subject could perform the entire exercise without any therapist intervention. During the training phase, two subjects did not understand how to manually choose/switch the sphere to try as the color of the button used to switch the sphere remained constant without matching the color of the sphere to squeeze (as in the testing phase) and at the same time, with only five available colors in the pushbutton keyboard, overlapped with the color of one sphere in the difficulty levels with five objects. One subject did not understand how to repeat the training phase. During the testing phase, only four subjects learned how to catch and identify the falling halos, due to difficulties in understanding the catching strategy (six out of ten subjects), controlling and maintaining the grip aperture during catching or squeezing (three out of ten), perceiving the stiffness differences (five out of ten). The median performance (i.e., number of spheres caught and identified versus total number of spheres) of the subjects within the three blocks was 27.00(16.38-31.50)%.

### 3.5 Additional Spontaneous Feedback

During the trial, the subjects reported additional spontaneous feedback. Three subjects recommended to modify the elbow support of the RHK. They asked to simplify the adjustment of the elbow support height with respect to the finger pads, which is currently done manually with two levers, and to constrain the forearm to the elbow support with straps to avoid large elbow movements during active pronosupination tasks (e.g., in the tunnel exercise). Two subjects reported mild/moderate pain in the fingers due to the finger straps, which were tightened to avoid finger slippage out of the thin handle surface. It was also reported that such finger pads might not allow a subject with high motor impairment to accurately control and perceive finger forces, as the contact between fingers and finger pads occurred only at the fingertips. Finally, the supervising therapist reported that to increase the safety of the device, all the mechanical parts of the robot (e.g., mechanical transmissions) that could get in contact with the user should be covered to avoid snag hazards (e.g., of the fingers).

## 4 Discussion

This paper presented the design and rigorous preliminary usability evaluation of a therapy platform (i.e., end-effector haptic device with new physical and graphical user interfaces and two novel therapy exercises) that aims to enable minimally-supervised robot-assisted therapy of hand function after stroke. This approach promises to be a suitable solution to increase the therapy dose offered to subjects after stroke either in the clinic (e.g., by allowing the training of multiple subjects in parallel, or additional training during the subject’s spare time in an unsupervised robotic gym), or at home after discharge, with the potential to maximize and maintain long-term therapy outcomes. A careful and quantitative pilot usability evaluation allows to preliminarily assess if the platform could be applicable in minimally-supervised conditions and which modifications are necessary to increase the feasibility of this therapy approach in a real-world minimally supervised scenario (e.g., in the clinic).

### 4.2 Minimally-Supervised Therapy is Possible upon Short-Term Exposure

Through the development of a modular graphical user interface and novel therapy exercises, we proposed a subject-tailored functional therapy platform that could be used upon first exposure by subjects after stroke in a single session with minimal therapist supervision. The platform was developed to meet a tradeoff between different requirements, namely to provide active task-oriented exercises similar to conventional exercises (Ranzani et al., 2020), while guaranteeing ease of use and subject compliance to the therapy program (motivation) while requiring minimal supervision both from a clinical (therapist) and technical (operator of the device) point of view (Zajc and Russold, 2019). Particular attention was dedicated to the optimization of the virtual reality interfaces, both in the graphical user interface and in the exercises to be easily usable/learnable, efficient (in terms of workload) and motivating, since it was shown that enriched virtual reality feedback might facilitate an increase in therapy dose and consequent improvements in arm function (Laut et al., 2015; Laver et al., 2015; Johnson et al., 2020).

In clinical settings, minimally-supervised therapy has rarely been investigated and documented (Büsching et al., 2018; McCabe et al., 2019). Starting from Cordo et al. 2009, few robotic devices have been proposed for minimally-supervised upper limb therapy at home (Cordo et al., 2009; Zhang et al., 2011; Lemmens et al., 2014; Sivan et al., 2014; Wolf et al., 2015; Hyakutake et al., 2019; McCabe et al., 2019). Only one device includes a virtual reality interface to increase subject motivation, and most of these devices only provide basic adaptive algorithms to customize the therapy plan to the subject needs. The device settings (e.g., lengths, sizes, finger pads) and exercise parameters are mostly manually tuned by the therapist at the beginning of the therapy protocol, while the subject performance is either ignored, telemonitored, or minimally-supervised by the therapist.

Typically, these devices were evaluated in research settings in terms of clinical efficacy, but they lacked usability evaluations, which would have been more informative and better correlated with their real-world adoption in a minimally-supervised scenario (Turchetti et al., 2014), as well as with their safety and feasibility. To obtain meaningful usability results that can be transferred to real-world use, therapy goals and use environment should be precisely defined.

### 4.2 The Platform was Attributed High Usability, with Suggestions for Minor Improvements

We achieved very positive usability results with our therapy platform, with system usability scores between good and excellent (i.e., between 70 and 90 out of 100) for the user interface and tunnel exercise, between OK and good (i.e., between 50 and 80 out of 100) for the sphere exercise, as well as TLX scores within the target workload boundaries. This study revealed that, even with only few minutes of instruction, it is easy to learn how to use the platform, use the colored pushbutton keyboard, navigate through the graphical user interface, insert the hand into the device finger pads and perform the exercises (i.e. particularly the tunnel exercise). Overall, the time needed to learn how to use the platform (instruction and training phase) and perform the experiment with minimal supervision (experimental phase) seems adequate for a first use of the platform (i.e., less than 60min), meaning that future users could be introduced to the platform within a single to two therapy sessions. The usability results showed an inverse trend with age but not with the impairment level of the subjects (either global, distal, or related to manual dexterity), reaching worst results for the oldest subject (age 87). However, this result is not significant, particularly with our small user sample size.

Usability evaluations of rehabilitation robots reported in literature yielded scores between 36 and 90 out of 100 using different usability questionnaires, including the SUS (Pei et al., 2017), “Cognitive Walkthrough” and “Think Aloud” methods (Valdés et al., 2014), custom-made questionnaires or checklists (Chen et al., 2015; Smith, 2015), which can be based on the Technology Acceptance Model (Davis, 1985). Only Sivan et al. 2014 proposed a basic evaluation of the usability of a minimally-supervised robotic platform considering the time needed to learn how to use the device independently and the total time of use of the device, and based on the experiments and user feedback identified design aspects that could be improved (Sivan et al., 2014). Human-centered designs based on usability evaluations have become best-practice in the medical field in recent years (Wiklund and Wilcox, 2005; Oviatt, 2006; Shah and Robinson, 2006; Blanco et al., 2016; ISO 9241, 2019), but the elicitation of standardized usability requirements and evaluations is still particularly challenging due to the heterogeneity of user groups, needs and environments (e.g., clinic or home) (Shah and Robinson, 2006), and the small sample sizes typically considered in this type of studies (van Ommeren et al., 2018). Therefore, it is generally difficult to compare the usability evaluations among different platforms.

The usability evaluation proposed in this work allows to quantify different aspects of usability, such as platform usability and learnability, perceived workload for the user, and ability to independently perform the tasks required during a minimally-supervised use of the platform. The early identification of these aspects during the development of the platform will allow us to improve design points that could bias the clinical applicability and testing of the platform with subjects. For instance, through our detailed usability analysis we highlighted key aspects regarding the physical and graphical user interfaces (e.g., handle size and shape, as well as button shape and color coding), and the exercise architecture (e.g., closer to activities of daily living). These aspects would not affect the feasibility of using the platform in clinical settings but would certainly impact its adoption in minimally-supervised settings.

### 4.3 Therapy Exercises are Functional, Motivating and Respect Target Workload Levels

The mental, physical and effort workloads in the exercises were rather high while frustration and performance workloads were rather low. The overall usability was high for both exercises. This is apromising result, underlining that the two new functional/synchronous exercises are engaging without being overly frustrating. Improvements would be needed to slightly increase the mental/cognitive workload required in the tunnel exercise and the physical workload in the sphere exercise. In the latter, however, the task complexity should be slightly reduced, since the performance achieved in the exercise is still too low with respect to the target performance level (i.e., 70%) and requires a too high performance workload. Detailed descriptions of minimally-supervised task-oriented exercises (e.g., requiring the functional training of multiple degrees of freedom) are rarely presented in literature (Lemmens et al., 2014; Sivan et al., 2014; Wolf et al., 2015). These exercises are often lacking engaging interfaces to enhance subject motivation (Zhang et al., 2011; McCabe et al., 2019) and are typically focused on pure motor training tasks, neglecting sensory and cognitive abilities. Based on the level of task complexity and on the usability results, our therapy exercises could be recommended for late stages of rehabilitation in a mildly/moderately impaired population. They could well complement the previously available exercises implemented on the rehabilitation robot that train either grasping or forearm pronosupination during passive proprioceptive tasks or active manipulation tasks (Metzger et al., 2014a).

### 4.4 The Platform is Safe and Could be Exploited For a Continuum of Robot-Assisted Care

After a guided instruction phase, our test tried to emulate a minimally-supervised environment in which the therapist intervened only in case help was required by the subject, as done in other studies performed in real-life minimally-supervised conditions (Lemmens et al., 2014; Sivan et al., 2014; Hyakutake et al., 2019). Throughout the test, the therapist intervention was needed on average less than 4 times per subject out of the 26 checklist items, mostly due to misunderstanding of the instructions or small software inconsistencies (e.g., unclear feedbacks, unclear color-function relations) without critical safety-related problems that would affect the applicability of the system with minimal supervision. These errors are expected to not occur anymore if the subjects were given a longer time for instructions and training. A continuum of use (over a larger time span) of our platform from supervised to minimally-supervised conditions would allow the user to familiarize with the system during the supervised sessions in the clinic and further continue the therapy seamlessly once the therapist is confident that the subject can safely train independently. Intervention minimization is useful to use the platform in the clinic, where a single therapist could supervise multiple subjects, and is essential in home environments, where external supervision is not always available or would require additional external communication channels (e.g., telerehabilitation (Wolf et al., 2015)). Safety and customization could be further increased through additional integrated robotic assessments. For example, two subjects reported a mild temporary increase in muscle tone, which can be physiologically induced by the active nature of the robotic assessments and exercises (Veerbeek et al., 2017). An increase in hand muscle tone may cause pain and negatively affect recovery, but could be monitored online throughout the therapy using robotic assessments incorporated into the therapy exercises (Ranzani et al., 2019).

### 4.5 Limitations

The results of this pilot study should be interpreted with respect to the relatively small sample size tested, which is, however, considered sufficient to identify the majority of the usability challenges (Virzi, 1992). The results reflect the usability of the platform for a mildly/moderately-impaired population in the chronic stage after stroke, which arguably is the target population for such a minimally-supervised therapy platform. The experiment lasted only one session, so it was not possible to evaluate how the subjects could learn to use the system in a longer term and in real-world minimally-supervised conditions, e.g. to assess if their motivation level would eventually drop after few sessions. Moreover, the scales proposed to evaluate the usability and workloads required by our platform can only partly capture the overall user experience, which should also account for user emotions, preferences, beliefs, physical and psychological responses before and after a longer use of the platform (Petrie and Bevan, 2009; ISO 9241, 2010; Meyer et al., 2019). Additionally, within this pilot study, it was not possible to implement the necessary usability adjustments that were identified and re-test the usability of the platform after modifications, but this should be assessed in the future. Finally, the presence of the technology developers during parts of the study (e.g., instruction and training phase) might have indirectly biased the usability evaluation performed by the subjects.

### 4.6 Future Directions

Future research should investigate how to equip rehabilitation robots with further intelligence to automatically propose therapy plans and settings based on objective measures, and provide comprehensive digital reports to remote therapists to monitor and document subjects’ progress. To be usable in a real-world minimally supervised scenario, the therapy platform would require minor adjustments identified throughout this study. Regarding the hardware, the finger pads should be wider to avoid finger slippage, and the pushbutton keyboard should include more buttons to allow consistent color-function and color-object mapping in GUI and exercises (e.g., insert one or two buttons uniquely for exercise control or quitting, to avoid color overlapping in difficulty levels requiring five objects/colors, such as in the sphere exercise). All the mechanical parts of the robot should be covered to avoid snag hazards, and an optical fingerprint reader could be added to the platform to simplify the access of multiple users to their therapy programs without the need to remember colored passwords. Regarding the software, based on the successful proof of concept with our new minimally-supervised exercises, the available assessment-driven supervised therapy exercises proposed by Metzger (Metzger et al., 2014b) will be redesigned to be usable with minimal supervision. As for the sphere exercise, attention should be devoted to the optimization of instructions/feedbacks clarity, and of task complexity and matching to real-world actions. The GUI should provide feedback to the therapist (e.g., subject performance and statistics) and possibilities to further customize the exercises (e.g., simplify graphical content for subjects with attention or cognitive deficits). Finally, a long-term study is required to evaluate the feasibility and usability of a continuum of robot-assisted care from supervised to minimally supervised conditions, and a mobile/portable device should be developed to allow the application of this approach also in the home environment.

## 4.7 Implications and Conclusions

The goal of this work was to develop and evaluate, in a single-session pilot study, the usability of a minimally-supervised therapy platform, allowing to perform functional, personalized and motivating task-oriented exercises at the level of the hand. Our findings demonstrate that a powered robot-assisted therapy device respecting usability and perceived workload requirements can be safely and intuitively used in a single session with minimal supervision by chronic stroke patients. This pilot evaluation allowed us to identify further design improvements needed to increase the platform usability and acceptance among the users. Our results open the possibility to use active robotic devices with minimal supervision to complement conventional therapies in real-world settings, offer increased dose with the existing resources, and create a continuum of care that progressively increases subject involvement and autonomy from the clinic to home.

## 6 Author Contributions

RR performed the data analysis and wrote the manuscript together with OL, JH and RG. LE and RR implemented the graphical user interface and the tunnel exercise. FV and RR implemented the sphere exercise. RR, OL, JH and RG defined the study protocol. JH was responsible for patient recruitment and screening. BE performed the experimental sessions with JH and RR.

## Data Availability

The discussion and conclusion of this work are based on data that are entirely reported within the manuscript. Any further detail will be provided by the authors upon request.

## 8 Acknowledgments

The authors would like to thank all the volunteers that participated in the pilot study, and Dominik G. Wyser and Jan T. Meyer for their valuable insights and discussions during the writing of this work.

